# Fecal genomic DNA extraction method impacts outcome of MinION based metagenome profile of tuberculosis patients

**DOI:** 10.1101/2021.11.15.21266154

**Authors:** Sukanya Sahu, Sandeep Rai Kaushik, Bidhan Goswami, Arunabha Dasgupta, Hritusree Guha, Ranjit Das, Sourav Saha, Anjan Das, Ranjan Kumar Nanda

## Abstract

In the present era, emergence of next generation sequencing approaches has revolutionized the field of gut microbiome study. However, the adopted DNA extraction step used in metagenomics experiments and its efficiency may play a critical role in their reproducibility and outcome. In this study, fecal samples from active and non-tuberculosis subjects (ATB/NTB, n=7) were used. Fecal samples of a subgroup of these subjects were subjected to Mechanical enzymatic lysis (MEL) and Phenol: Chloroform: Isoamyl Alcohol (PCIA) methods of DNA extraction and a third-generation sequencing platform i.e. MinION was employed for microbiome profiling. Findings of this study demonstrated that DNA extraction method significantly impacts the DNA yield and microbial diversity. Irrespective of the adopted method of DNA extraction, ATB patients showed altered microbial diversity compared to NTB controls. Also, the fecal microbial diversity details are better captured in samples processed by MEL method and may be suitable to be adopted for high-throughput gut microbiome studies.

## INTRODUCTION

Gut microbiota is composed of trillions of microorganisms belonging to diverse groups such as bacteria, virus and fungi.^1,2^ It also contributes and play a critical role in host metabolism, development and optimum function of the immune system to provide protection from pathogenic invasion.^3,4,5^ Profiling of this complex community brings useful information to understand the perturbed patho-physiology in diverse disease conditions.^6,7,8,9,10^ Identified targets may help to devise translatable solutions for optimum maintenance of gut homeostasis.^11,12,13,14,15^

In the past few decades, development of metagenomics tools promoted compositional and functional analyses of gut microbiota which were previously an unrealistic undertaking.^16,17^ A typical metagenomics study involves sampling, storage, processing, genomics data acquisition, informatics analysis and further validation. Adopted DNA extraction methods are known to influence the outcome of metagenomics experiments.^18,19^ Therefore, it is important to select appropriate DNA extraction methods to explore diverse microbial communities including the minor contributors.

In this pilot scale study, we aimed to capture the gut microbial diversity of active tuberculosis (ATB) and non-tuberculosis (NTB) subjects. The influence of the adopted DNA extraction method on the microbial diversity in a subset of samples processed by Mechanical Enzymatic Lysis (MEL) and Phenol: Chloroform: Isoamyl Alcohol (PCIA) methods were also monitored. Findings of this study may be useful to get reproducible inter- and intra-laboratory gut microbial profiling results by improving the coverage in different disease conditions.

## METHODS

### Ethics statement

This study is part of an ongoing project approved by the Institute Human Ethics Committees of the Agartala Government Medical College, Agartala (protocolF.4[6-9]/AGMC/Academic/IEC Committee/2015/8965, dated 25 April 2018) and International Centre for Genetic Engineering and Biotechnology, New Delhi (ICGEB/IEC/2017/07).

### Subject recruitment and sample collection

Subjects presenting with symptoms of cough (> 2 weeks), fever, weight loss and night sweat to the outpatient department of the clinical site were recruited after receiving signed informed consent. Subjects (n=7, male/female: 4/3; age 40.9 (30-57) in years) of both gender and above 18 years of age were included. Epidemiological details of these study subjects are presented in Table 1. Collected sputum samples (∼5 ml) were subjected to acid fast bacilli microscopy by Ziehl–Neelsen staining, and cartridge based nucleic acid amplification test (GeneXpert). Subjects with all positive test results grouped as active tuberculosis (ATB) and negative test results as non-tuberculosis (NTB). These NTB subjects were suffering from other respiratory disease conditions like pneumonia or lung cancer or asthma or chronic obstructive pulmonary disease (COPD) as established by clinical decisions (Figure 1). Fecal samples (∼ 50 grams) were collected by the study subject using a sterile container without touching the surroundings. Within 60 mins of collection, fecal samples were stored at -80°C till further analysis.

**Table1.** Epidemiological details of the study subjects

| Demographic details | Total | Active tuberculosis (ATB) | Non-tuberculosis (NTB) |
| --- | --- | --- | --- |
| Population size | 7 | 4 | 3 |
| Mean age (range) in years | 40.9 (30-57) | 41 (32-57) | 40.7 (30-50) |
| Gender (Female/Male) | 3/4 | 1/3 | 2/1 |
| Body mass index (kg/m<sup>2</sup>) | 17.08 | 17.70 | 15.84 |
| Cough (y/n) | 7/- | 4/- | 3/- |
| Expectoration (y/n) | 6/1 | 4/- | 2/1 |
| Haemoptysis (y/n) | -/7 | -/4 | -/3 |
| Chest pain (y/n) | 4/3 | 2/2 | 2/1 |
| Breathlessness (y/n) | 1/6 | 1/3 | -/3 |
| Wheeze (y/n) | -/7 | -/4 | -/3 |
| Body temp. (>37°C/normal) | 2/5 | 1/3 | 1/2 |
| Smoking (y/n) | 4/3 | 3/1 | 1/2 |
| Alcoholism (y/n) | 4/3 | 3/1 | 1/2 |
| Sputum AFB Test (+/-ve) | 3/4 | 3/1 | -/3 |
| GeneXpert test result (+/-ve) | 4/3 | 4/- | -/3 |

**Figure 1:**
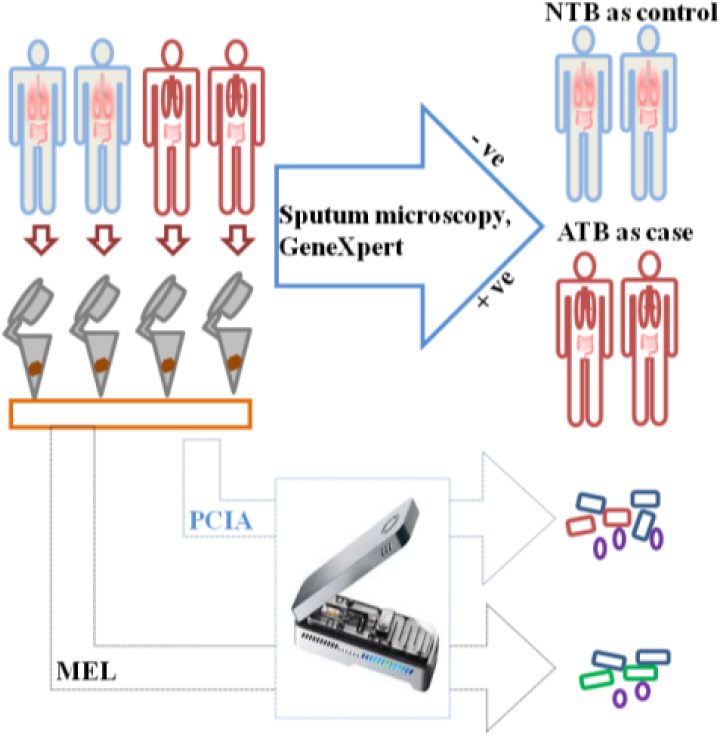
Study subject classification for fecal sample collection, processing and microbiome analysis using MinION. Study subjects with positive sputum microscopy and GeneXpert results were grouped as case (active tuberculosis: ATB) and with both negative results were grouped as non-tuberculosis (NTB) control groups. Methods (mechanical-enzymatic lysis method: MEL and Phenol: Chloroform: Isoamyl Alcohol: PCIA) adopted for fecal genomic material isolation.

### Genetic material extraction

Two different methods (Mechanical-Enzymatic Lysis method: MEL and Phenol: Chloroform: Isoamyl Alcohol method: PCIA) were used for genomic DNA extraction from fecal samples of the study subjects. The steps involved in both the methods are schematically presented in Figure 2.

**Figure 2:**
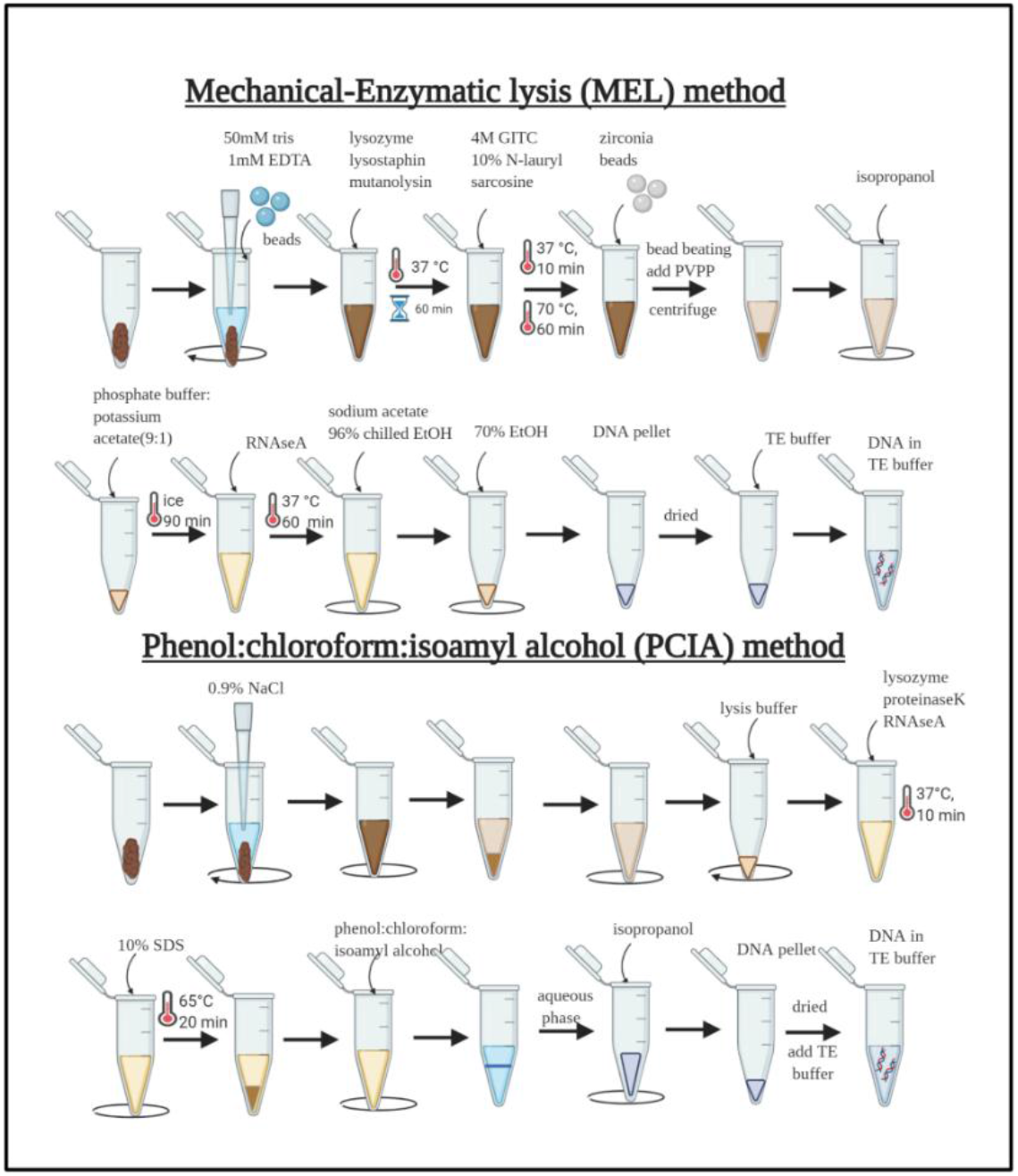
Schematic presentation of the steps involved in the adopted fecal genomic DNA extraction methods (mechanical-enzymatic lysis method: MEL and Phenol: Chloroform: Isoamyl Alcohol: PCIA) used in this study. This image was generated using BioRender Software (**http://www.biorender.com/**).

### MEL method of fecal sample processing

Fecal samples were brought to room temperature and equal amount (150 mg) of it was transferred to pre-chilled micro-centrifuge tube and mixed with TE buffer (50 mM Tris-1mM EDTA, pH 8.0, 200 µl). To it, glass beads (n=4, 2.7 mm) were added and vortexed (∼45 sec). After removal of glass beads, enzyme solution mix (lysozyme (50 µl, 10 mg/ml), mutanolysin (6 µl, 2 KU/ml) and lysostaphin (3 µl, 4 KU/ml)) were added and incubated at 37° C for 1 hour. To the reaction mixture, Guanidinium thiocyanate (250 µl, 4M) was added and mixed gently for 45 seconds before adding N-lauryl sarcosine (300 µl, 10 %) and incubated at 37° C for 10 minutes at 300 rpm in a thermomixer. After vortexing, samples were centrifuged at 16,000 *g* for a few seconds and incubated at 70°C for 1 hour. After adding Zirconia beads (300 mg, 0.1 mm) to the samples, bead beating was carried out in two cycles (30 sec each) for 2 minutes. After adding PVPP (15 mg), the sample mixture was vortexed before centrifuging at 16,000 *g* for 3 min at room temperature. The supernatant was transferred to a fresh tube and the pellet was washed twice with Tris (50 mM)-EDTA (20 mM)-NaCl (100 mM)-PVPP (1%) solution (500 µl) and pooled. To it, Isopropanol (2 ml) was added and after mixing for a few minutes, samples were incubated at room temperature for 10 minutes before centrifuging at 16,000 *g* for 10 minutes. The pellet was collected, dried at room temperature and phosphate buffer: potassium acetate (1 ml, 9:1) was added before incubating on ice for 90 minutes (or -20°C overnight). The sample was centrifuged at 16,000 *g* for 30 minutes at 4°C and to the supernatant, RNaseA (4 µl, 10 mg/ml) was added and incubated at 37°C for 60 minutes. Sodium acetate (3M, pH 5.2, 50 µl) was added to each tube followed by ice-cold ethanol (1 ml, 96 %) and mixed by inverting the tubes a few times before incubating at room temperature for 5 minutes. After centrifuging at 16,000 *g* for 15 minutes at 4 °C, the supernatant was discarded followed by washing the pellet twice with ice-cold ethanol (70%). The dried pellet was resuspended in Tris-EDTA buffer (200 µl, 10 mM-1 mM, pH 8.0) and extracted genomic DNA amount was measured using Qubit 3.0 fluorometer (Q33216, Thermo Fischer Scientific, United States).

### PCIA method of fecal sample processing

To fecal samples (100 mg each, after bringing to room temperature), normal saline solution (1 ml) was added and centrifuged at 645 *g* at room temperature for 2 minutes to collect the supernatant and centrifuged again at 7,168 *g* at room temperature for 1 min. The microbial pellet was resuspended with PBS (pH 7.4, 1 ml) and centrifuged at 645 *g* for 2 min at room temperature and the resulting supernatant was centrifuged at 7,168 *g* for 1 min at room temperature. To the microbial pellet, lysis buffer (500 µl) and proteinase K (2 µl, 20 mg/ml) was added and incubated for 10 min at 37°C with gentle shaking at 300 rpm. To the reaction mixture, SDS (50 µl, 10 %) was added and incubated at 65° C for 20 min. The mixture was centrifuged at 12,114 *g* for 5 min at room temperature and to the supernatant, an equal volume of Phenol: Chloroform: Isoamyl Alcohol (25:24:1) was added. The reaction mixture was centrifuged at 7,168 *g* for 5 min at room temperature. The aqueous phase was collected and isopropanol (0.6 volume) was added before centrifuging at 12,114 *g* for 5 min at room temperature. To the DNA pellet, ethanol (ice-cold, 70 %, 1 ml) was added before centrifuging at 7,168 *g* for 5 min at room temperature. The dried pellet was re-suspended in TE buffer (pH 8.0, 50 µl) and extracted genomic DNA amount was quantified using Qubit 3.0 fluorometer (Q33216, Thermo Fischer Scientific, United States).

### Library preparation

Isolated genomic DNA (∼700 ng) from faecal samples of all study subjects belonging to ATB and NTB groups were taken for library preparation. Nuclease free water (12931S, NEB, United States) was added to each DNA sample to a final volume of 50 µl, followed by addition of end repair buffer (E7546S, NEB, 7 µl), end repair enzyme (E7546S, NEB, 3 µl) and incubated at 20° C for 5 minutes followed by incubation at 65° C for 5 minutes. To the AMPure beads (A63880, Beckman Coulter, United states, 60 µl), DNA prep was added slowly and incubated at room temperature for 5 minutes. These tubes were placed in a magnetic rack (NEB, United States) till the solution becomes clear (∼5 min). Carefully the supernatant was taken out without disturbing the pellet and washed twice with EtOH (70 %, 200 µl). These tubes were removed from the rack and nuclease free water was added (23.5 µl) and incubated at room temperature for 5 minutes. These tubes were kept on a magnetic rack for 5 minutes till the solution became clear and the supernatant was transferred to a separate micro-centrifuge tube. Barcodes (2.5 µl) were added to each tube followed by a blunt T/A ligase master mix (25 µl) and incubated at RT for 10 min. To this reaction mixture, AMPure XP beads (50 µl) were added and washed twice using ethanol (70 %). The pellet was resuspended with nuclease free water (NFW, 20 µl) and incubated at room temperature for 5 minutes and kept in a magnetic rack for another 5 minutes. Equal amounts of barcoded DNA from all samples were pooled in a tube (0.2 ml) and Barcode Adapter Mix (BAM, Oxford Nanopore, 20 µl) was added. Equal volume of Blunt/TA ligase (NEB, United States) was added to this reaction mixture and incubated for 20 minutes at 20° C. Then AMPure beads (0.4 times of the resultant volume) were added to the reaction mix and incubated at room temperature for 5 minutes and kept in a magnetic rack for 5 minutes. To the pellet, ABB Buffer (140 µl) was added, kept in a magnetic rack for 5 minutes and repeated twice. The tubes were removed from the magnetic rack, using an elution buffer (16 µl) the pellet was resuspended and incubated at room temperature for 5 minutes followed by keeping them on a magnetic rack for another 5 minutes and the supernatant was transferred to a clean tube.

### Priming

Flow cell priming mix was prepared by mixing Running Buffer with Fuel Mix (RBF, 576 µl) and nuclease free water (624 µl). The priming port was opened in the flow cell (FLO-MIN107) 9.4.1 and after removing the buffer (20-30 µl), priming mix (800 µl) was loaded into the flow cell through the priming port by rotating the pipette anticlockwise. Then the flow cell was kept as such for 5-10 minutes.

### Library loading

Before loading, the library was prepared by adding Library Loading Beads (LLB, 25.5 µl) and Running Buffer with Fuel Mix (RBF, 35 µl) to the pooled library to a final volume (75 µl). The priming mix (200 µl) was added to the flow cell through the priming port. Then the prepared library (75 µl) was loaded into SpotON by drop by drop. Then the SpotON sample port cover was gently replaced followed by closing the priming port and replacing the MinION lid.

### Sequence data processing

Upon completion of sequencing run, the reads generated in FAST5 format were demultiplexed and converted to FASTQ using Albacore software with barcoding option. Base calling and analysis of the sequencing reads was performed in real-time using EPI2ME, a cloud-based analysis platform of Oxford Nanopore (Oxford Nanopore Technologies Limited, UK).

### Statistical analysis

Microbiota profiles generated after nanopore sequencing were plotted and visualized using Microsoft Excel 2019. Bland-Altman plots were made to explore the differences in the microbiota profiles generated by nanopore sequencing using two different methods for top seven prevalent genera. Venn diagrams were plotted using a web tool (http://bioinformatics.psb.ugent.be/webtools/Venn/).

## RESULTS

### Tuberculosis patients showed altered gut microbiome profiles

Genetic material extracted from fecal samples of ATB and NTB subjects using PCIA method and metagenomics profiling by MinION platform showed disease specific differences. In majority of the study samples irrespective of ATB or NTB groups, *Bacteroides, Bifidobacterium, Escherichia, Enterococcus, Lachnoclostridium, Parabacteroides* and *Fecalibacterium* genera were commonly detected. Overall, *Bacteroides* (33%) was found to be most abundant followed by *Escherichia* (13.3%) and *Enterococcus* (12.5%) (Figure 3A). Taxonomic profile of identified bacterial genera in both the ATB and NTB study subjects showed quantitative variation (Figure 3B). Higher abundance of *Enterococcus* in ATB (1.5-62.2%) compared to NTB subjects (<5.5%) was observed. Whereas higher *Bacteroides* abundance was observed in NTB subjects (40-79.8%) than ATB (6-37.4%). Higher abundance of *Lachnoclostridium* and *Bifidobacterium* were observed in ATB subjects than in NTB subjects. In one out of 4 ATB study subjects, *Akkermansia* (21.2%) and in a single NTB subject *Acidaminococcus* (5.8%) were identified.

**Figure 3:**
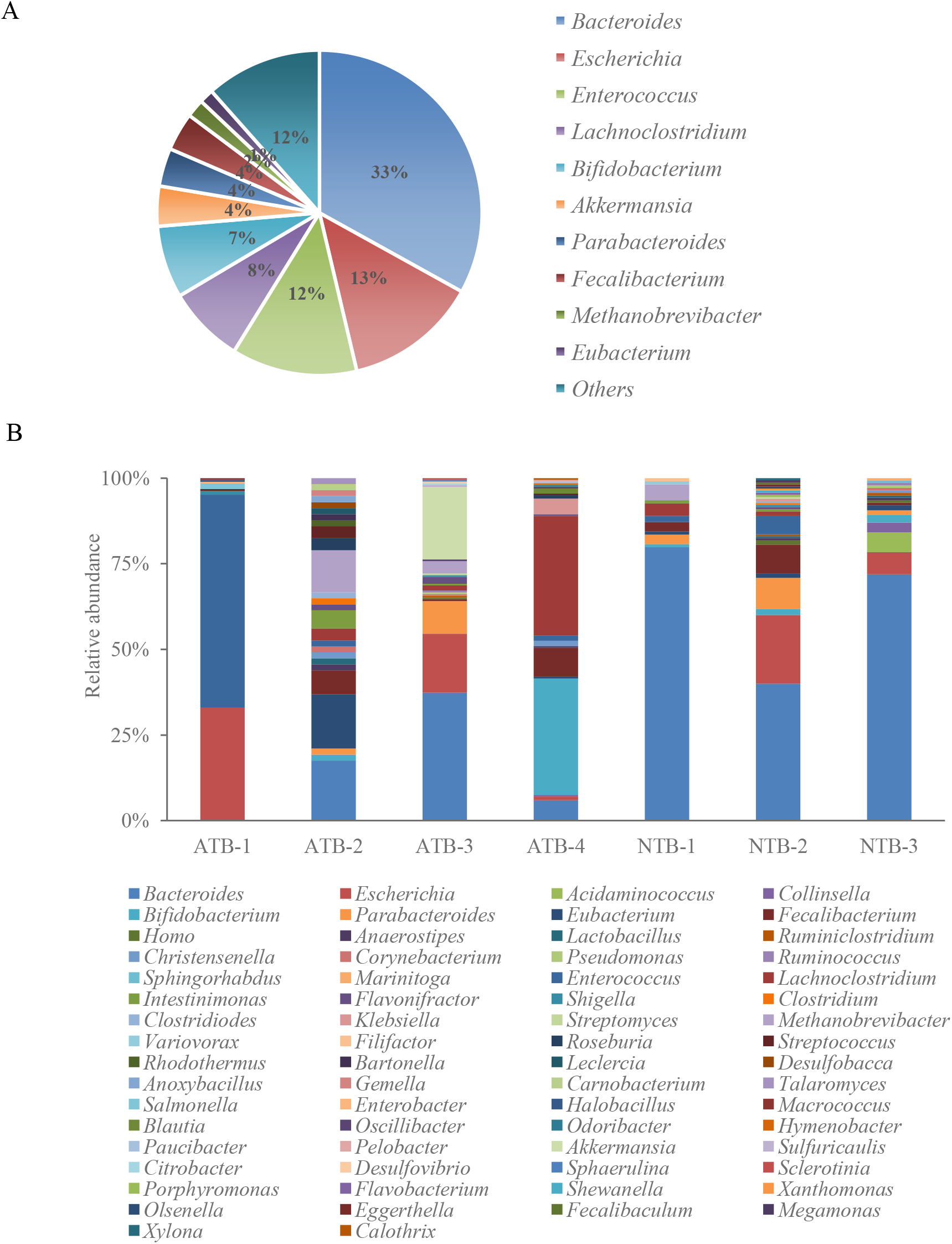
Microbiota composition of ATB and NTB subjects processed using Phenol: Chloroform: Isoamyl Alcohol (PCIA) PCIA method. A. Pie chart showing the overall distribution of bacterial genera in ATB and NTB subjects. B. Taxonomic profile of bacterial genera identified in the gut microbiome of ATB and NTB subjects.

### Gut microbiome profiling of ATB and NTB subjects processed by MEL and PCIA methods

To monitor the contribution of DNA extraction methods on outcome of metagenomics study, fecal samples (n=2, ATB/NTB:2/2) were processed using two different genomic DNA extraction methods. The total genomic DNA yield from the study samples processed using the MEL (18.87-112.23 ng/mg, n=4) method was tenfold higher than PCIA (1.18-9.54 ng/mg, n=4) (Figure 4A).

**Figure 4:**
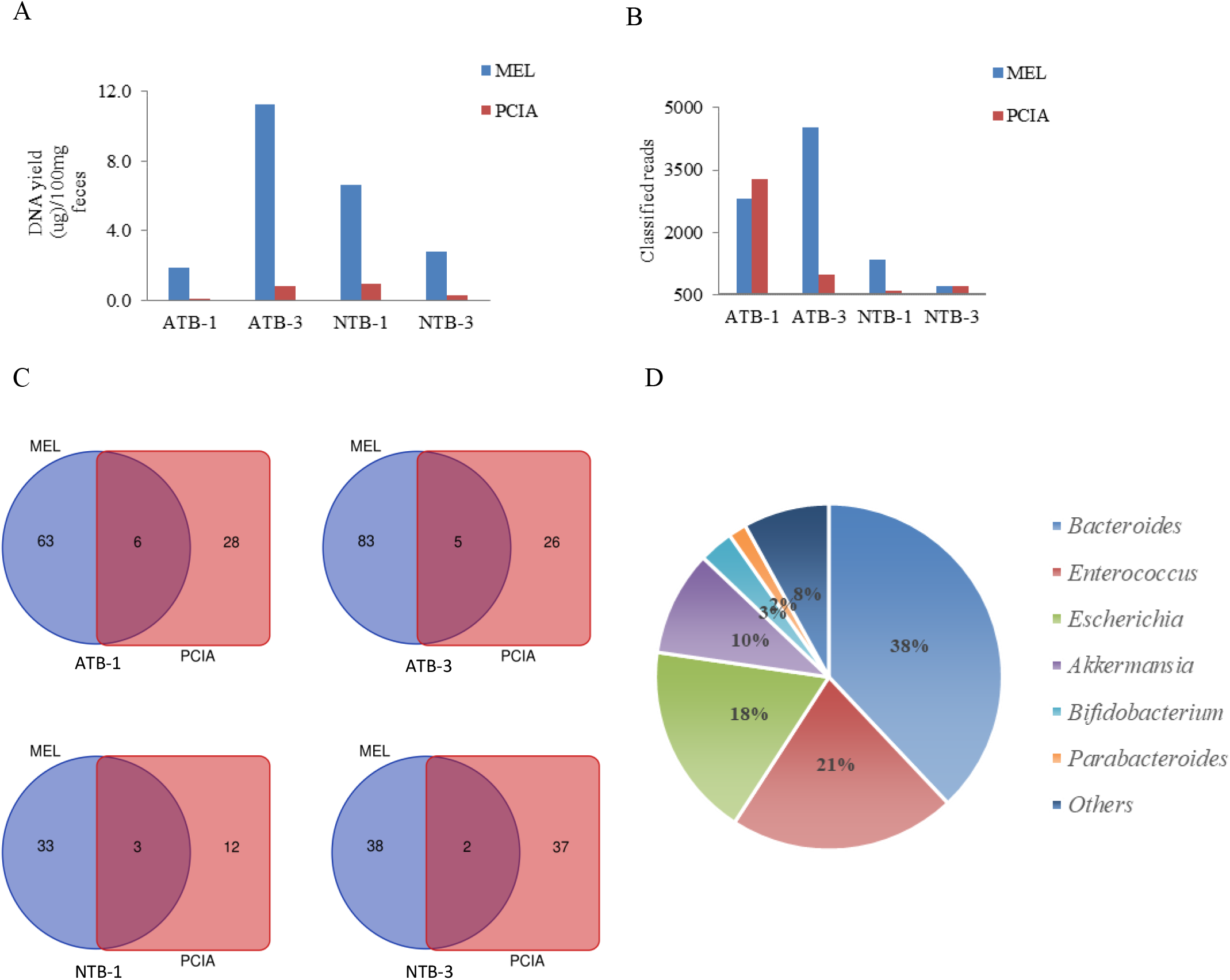
Effect of Mechanical-enzymatic lysis (MEL) and Phenol: Chloroform:Isoamyl Alcohol (PCIA) methods of DNA extraction on microbiome composition. A. Total fecal genomic DNA yield was found to be higher when MEL method was used. B. Total number of classified reads generated from the genomic DNA of fecal samples from drug naive active tuberculosis and non-tuberculosis cases extracted using MEL and PCIA method and analyzed using MinION platform. C. Venn diagrams representing the core unique and shared gut microbiomes at genus levels in processed samples. D. Pie chart showing overall distribution of bacterial genera in MEL and PCIA methods.

Upon analysis using MinION sequencing, a higher number of reads were identified in the majority of these samples processed using the MEL method (Figure 4B). Total number of taxa identified from the samples processed using MEL and PCIA methods showed limited overlap (<6.5%, n=4) (Figure 4C). *Bacteroides* (38%), *Enterococcus* (21.2%), *Escherichia* (18.1%), *Bifidobacterium* (3.2%), *Akkermansia* (9.8%), *Parabacteroides* (1.7%), Collinsela (0.7%), *Fecalibacterium* (0.5%) and *Lachnoclostridium* (0.5%) were the most prevalent genera captured by both these methods (Figure 4D). *Bacteroides, Escherichia and Bifidobacterium* were the most prevalent genera in samples processed using either MEL or PCIA method and identified in the majority of these samples. Whereas *Akkermansia* and *Parabacteroides* were detected in one out of four samples processed using PCIA and in all samples processed using MEL method. The overall abundance of *Enterococcus, Bifidobacterium, Akkermansia, Parabacteroides, Collinsella* and *Lactobacillus* were higher in samples processed using the MEL method as compared to PCIA. Higher *Bacteroides* abundance was observed in samples processed using PCIA (49.4-94.4%) than MEL method (23.5-67.3%). Fecal samples processed using MEL yielded higher bacterial diversity at genus levels including identification of unique genus such as *Penibacillus, Ruminoclostridium, Blautia, Ornithobacterium, Prevotella* as compared to PCIA. Adopted DNA extraction methods influenced the outcome of metagenomic profiles of the same samples irrespective of their group and MEL method captures better microbial diversity (Figure 5).

**Figure 5.**
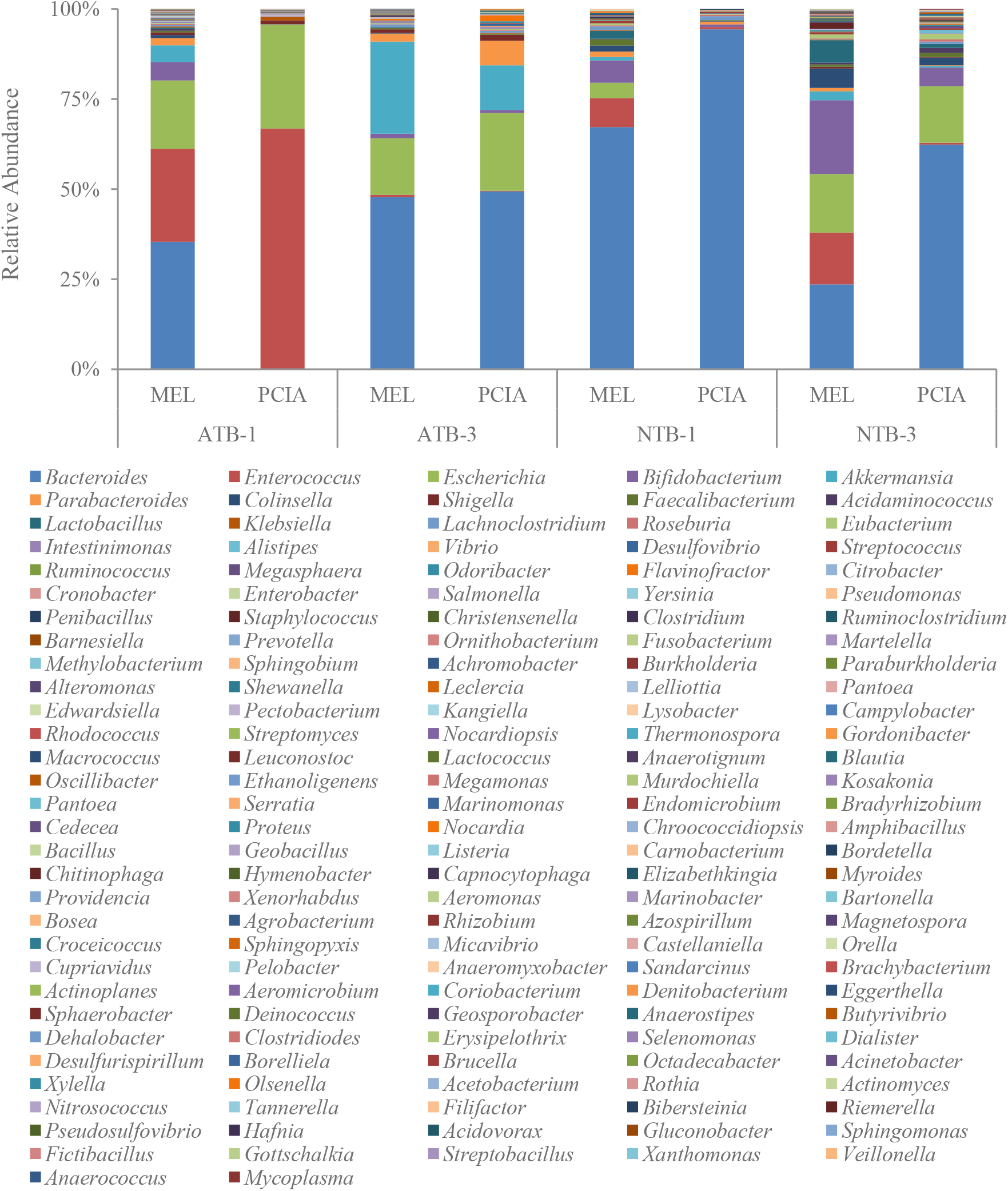
Taxonomic profile of bacterial genera identified in the gut microbiome of ATB and NTB subjects showed DNA extraction method specific differences. Fecal samples were processed using Mechanical-enzymatic lysis (MEL) and Phenol: Chloroform: Isoamyl Alcohol (PCIA) methods for DNA extraction.

Further, the agreement between MEL and PCIA methods was evaluated for the top seven contributing genera (*Bacteroides, Enterococcus, Bifidobacterium, Akkermansia, Parabacteroides, Lactobacillus* and *Collinsella)* and Bland-Altman plots showed method specific differences (Figure 6A-H). Certain degree of biasness to the MEL method in detecting *Enterococcus* and *Akkermansia* was observed. The wide confidence intervals and higher mean differences variation are expected in studies involving small sample size (Figure 6).

**Figure 6:**
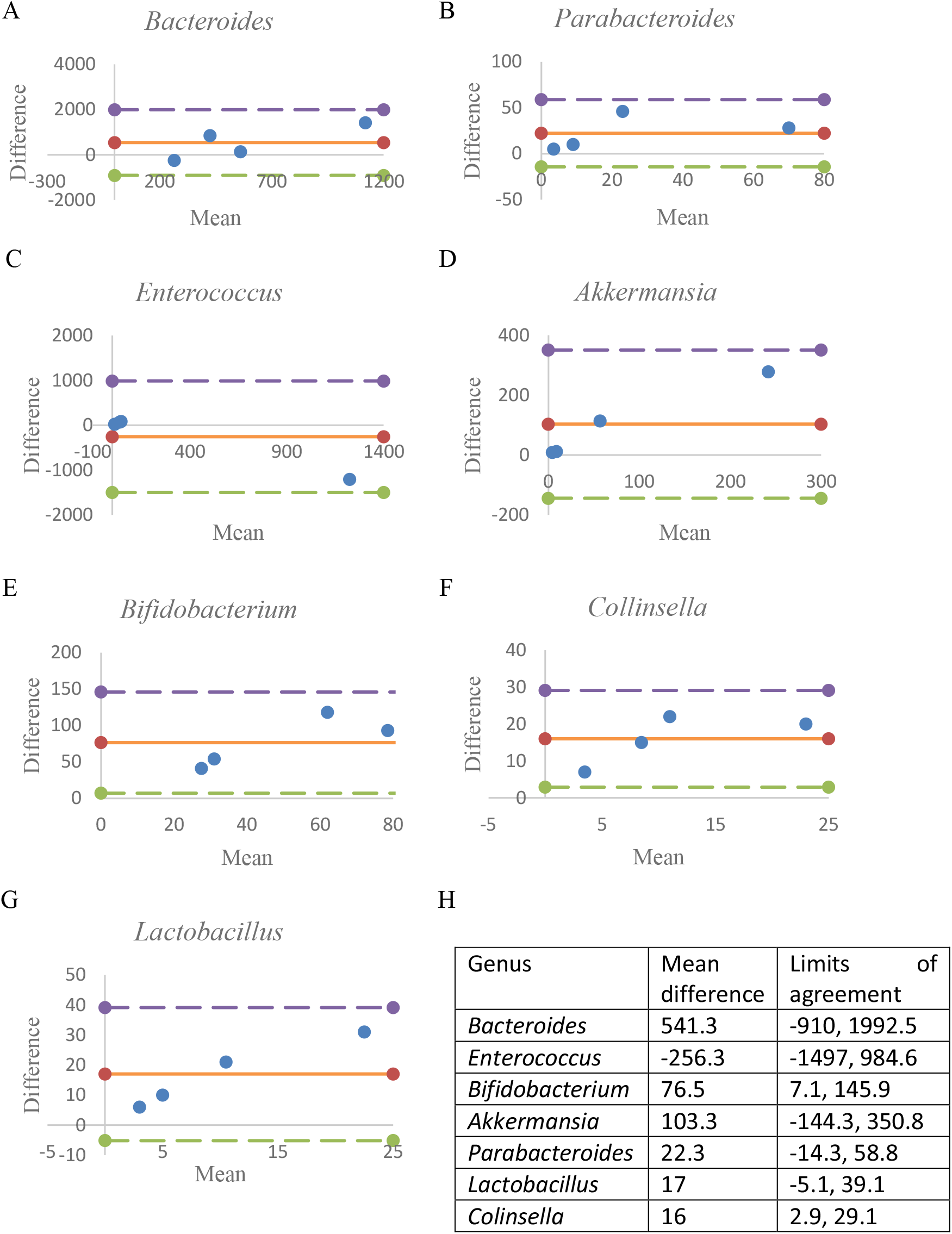
Bland-Altman plots of seven main genera present in the gut microbiota. Bland-Altman plots were generated for the seven main genera: (A) Bacteroides (B) Parabacteroides (C) Entereococcus (D) Akkermansia (E) Bifidobacterium (F) Colinsella and (G) Lactobacillus. For each genus, the mean difference between the two extraction methods (MEL vs PCIA) and the limits of agreement (95% reference interval) were calculated and shown (H).

## DISCUSSION

Metagenomics studies capture detailed microbial diversity in varied samples and useful for a better understanding of the pathophysiology in healthy and perturbed conditions.^20,21^ In fact, in certain diseases, supplementation with probiotics provided a positive outcome in resolving the perturbed conditions.^22^ However, it involves multiple critical steps such as sampling, storage, processing, sequencing and informatics analyses which can impact the outcome affecting intra- and inter-laboratory reproducibility.

Several reports including the MicroBiome Quality Control project (MBQC) and the International Human Microbiome Standards (IHMS) group highlighted majority of experimental variability to the adopted DNA extraction methods.^23,24^ As expected, fecal samples contain a diverse group of microbes and selecting an appropriate DNA extraction method might minimize the over- or under-representation of specific microbes. A third-generation sequencing platform i.e. MinION was used for profiling the extracted DNA, due to its portability, speed of data production, ease of use and ability to generate long reads in real time. And MinION and Illumina platforms generated comparable results from different biofluids including human nasal samples.^25^

Many metagenomics studies adopt commercial kits for DNA extraction and reports demonstrated that manual method processed fecal samples better captures the microbial diversity.^26^ In this study we compared two manual methods of DNA extraction for further processing. Commercial method of DNA extraction has its own advantage of intra and inter-laboratory reproducibility but if it captures partial diversity, it will impact the study outcome.

Like other disease conditions, infection also alters gut microbial diversity in humans. Few reports have demonstrated altered microbial diversity in pulmonary ATB patients with respect to NTB or healthy controls.^27,28^ In this pilot scale study, we employed the PCIA method of DNA extraction from ATB and NTB subjects and found differences in the gut microbiota composition between the groups. From the MinION results, higher abundance of *Lachnoclostridium, Enterococcus* and *Bifidobacterium* in the ATB subjects compared to NTB were observed and corroborated earlier findings.^29,30,31,32^

To further monitor the contribution and impact of the adopted method of DNA extraction, microbial diversity was monitored in fecal samples of ATB and NTB subjects processed using manual methods like MEL and PCIA, among other widely used commercial kits (QIAamp DNA stool MiniKit, Qiagen DNeasy kit, MoBio PowerFecal kit).^33^ The MEL method is a combination of enzymatic, mechanical and thermal treatment for disruption of bacterial cells and involves three different lytic enzymes namely lysozyme, lysostaphin and mutanolysin which targets both Gram-positive and Gram-negative bacterial cell wall, whereas PCIA uses enzymatic (lysozyme) and thermal treatment.^26,34^ Lysozyme hydrolyses 1,4-βglycosidic-linkages whereas lysostaphin acts on the polyglycine bridges present on the peptidoglycan layer of the cell wall and mutanolysin act against bacteria having O-acetylated peptidoglycan.^35,36,37,38,39,40^

The total genomic DNA yield was higher in MEL than the PCIA method corroborating earlier reports.^26,34^ It seems that lysozyme alone does not efficiently lyse the diverse bacterial cells and mechanical disruption by bead beating has higher DNA extraction efficiency.^41,42,43^ We observed higher DNA yield as well as better coverage on bacterial diversity in the samples processed using MEL supporting earlier claims.^35,44,45,46^ In this comparative analysis, majority of identified taxa belonged to the class Bacteroidia, Gammaproteobacteria, Bacilli, Actinobacteria, Verrucomicrobiae,Methanobacteria and Clostridia irrespective of the adopted extraction methods. Interestingly, the relative abundances of these classes were higher in the majority of the samples processed using the MEL method as compared to PCIA. Flavobacteria class was identified in the majority of these samples processed using the MEL method. The gram-positive bacterial class Bacilli, Actinobacteria, Clostridia were more abundant along with the gram-negative bacteria class Bacteroidia, Gammaproteobacteria, Verrucomicrobiae in samples processed using MEL method compared to those extracted using PCIA.

The difference between the ATB and NTB subjects, if any, used for comparison of DNA extraction efficiency among the two methods was also explored. Higher abundance of the Bacteroidia class was observed in the ATB subjects as compared to NTB samples processed using MEL method corroborating earlier reports.^47^ We observed *Bifidobacterium* abundance associated with NTB controls whereas *Faecalibacterium* and *Roseburia* were enriched in ATB patients as reported earlier.^48^ In addition, the bacterial genus *Enterococcus, Akkermansia* and *Parabacteroides* were enriched in the TB patients. *Prevotella* and *Eubacterium* levels didn’t show any differences between ATB and NTB groups unlike reported earlier.

Few limitations of this study include the small sample size used for these pilot scale comparative experiments. However, the observations of this study corroborated earlier reports which showed that altered microbial diversity is influenced by the adopted DNA extraction methods. So, it is important to select an appropriate and efficient DNA extraction method for wider coverage of microbiome diversity to ensure reproducibility in microbiome profiling studies. Validation using complementary methods would be useful to substantiate these claims. Overall, in this study we demonstrated ATB subjects have different microbial diversity than NTB and DNA extraction method plays a critical role in capturing it.

## CONCLUSION

Microbiome profiling study and their outcomes depend on several factors including adopted methods of sample collection, storage, processing steps, sequencing platforms used for data acquisition and bioinformatics tools used for such analysis. Our findings suggest that the MEL method yields a higher amount of DNA and MinIOn based data showed improved coverage of microbial diversity details than the PCIA method. Thus, the MEL method may be used as a preferred method to profile the differences between perturbed and control subjects for a better understanding of disease and underlying conditions.

## Data Availability

All data produced in the present study are available upon reasonable request to the authors.

## ACKNOWLEDGEMENTS

We would like to acknowledge the contribution of members from the clinical site used for patient recruitment and sample collection. Funding from Department of Biotechnology (DBT), Government of India for supporting activities through research grant (MDR-TB/2017/39) and core support from International Centre for Genetic Engineering and Biotechnology, New Delhi to RKN is highly acknowledged. SS is supported by fellowship from Council of Scientific and Industrial Research, Government of India. SRK is supported by Department of Biotechnology, Government of India.

## AUTHORSHIP CONTRIBUTIONS

SS and SRK conducted the experiments. SS and RKN drafted the manuscript taking inputs from all co-authors. Recruitment of study subjects and classification was conducted by HG, RD, SS under the guidance of AD, BG and AD. Funds for this work was generated by RKN and AD.

## Notes

### Competing Interest Statement

The authors have declared no competing interest.

## REFERENCES

1. Bull, M.J. and Plummer, N.T., 2014. Part 1: The human gut microbiome in health and disease. Integrative Medicine: A Clinician’s Journal, 13(6), p.17.

2. Rath, C.M. and Dorrestein, P.C., 2012. The bacterial chemical repertoire mediates metabolic exchange within gut microbiomes. Current opinion in microbiology, 15(2), pp.147–154.

3. Guinane, C.M. and Cotter, P.D., 2013. Role of the gut microbiota in health and chronic gastrointestinal disease: understanding a hidden metabolic organ. Therapeutic advances in gastroenterology, 6(4), pp.295–308.

4. O’Hara, A.M. and Shanahan, F., 2006. The gut flora as a forgotten organ. EMBO reports, 7(7), pp.688–693.

5. Sekirov, I., Russell, S.L., Antunes, L.C.M. and Finlay, B.B., 2010. Gut microbiota in health and disease. Physiological reviews.

6. Yaung, S.J., Church, G.M. and Wang, H.H., 2014. Recent progress in engineering human-associated microbiomes. Engineering and analyzing multicellular systems, pp.3–25.

7. Sartor, R.B., 2014. The intestinal microbiota in inflammatory bowel diseases. In Nutrition, Gut Microbiota and Immunity: Therapeutic Targets for IBD (Vol. 79, pp. 29–39). Karger Publishers.

8. Becker, C., Neurath, M.F. and Wirtz, S., 2015. The intestinal microbiota in inflammatory bowel disease. ILAR journal, 56(2), pp.192–204.

9. Ley, R.E., Turnbaugh, P.J., Klein, S. and Gordon, J.I., 2006. Human gut microbes associated with obesity. Nature, 444(7122), pp.1022–1023.

10. Round, J.L., O’Connell, R.M. and Mazmanian, S.K., 2010. Coordination of tolerogenic immune responses by the commensal microbiota. Journal of autoimmunity, 34(3), pp.J220–J225.

11. Venkataraman, A., Sieber, J.R., Schmidt, A.W., Waldron, C., Theis, K.R. and Schmidt, T.M., 2016. Variable responses of human microbiomes to dietary supplementation with resistant starch. Microbiome, 4(1), pp.1–9.

12. Martínez, I., Kim, J., Duffy, P.R., Schlegel, V.L. and Walter, J., 2010. Resistant starches types 2 and 4 have differential effects on the composition of the fecal microbiota in human subjects. PloS one, 5(11), p.e15046.

13. Walker, A.W., Ince, J., Duncan, S.H., Webster, L.M., Holtrop, G., Ze, X., Brown, D., Stares, M.D., Scott, P., Bergerat, A. and Louis, P., 2011. Dominant and diet-responsive groups of bacteria within the human colonic microbiota. The ISME journal, 5(2), pp.220–230.

14. Zhao, L., Zhang, F., Ding, X., Wu, G., Lam, Y.Y., Wang, X., Fu, H., Xue, X., Lu, C., Ma, J. and Yu, L., 2018. Gut bacteria selectively promoted by dietary fibers alleviate type 2 diabetes. Science, 359(6380), pp.1151–1156.

15. Chua, K.J., Kwok, W.C., Aggarwal, N., Sun, T. and Chang, M.W., 2017. Designer probiotics for the prevention and treatment of human diseases. Current Opinion in Chemical Biology, 40, pp.8–16.

16. Wang, W.L., Xu, S.Y., Ren, Z.G., Tao, L., Jiang, J.W. and Zheng, S.S., 2015. Application of metagenomics in the human gut microbiome. World journal of gastroenterology: WJG, 21(3), p.803.

17. Galloway-Peña, J. and Hanson, B., 2020. Tools for analysis of the microbiome. Digestive diseases and sciences, 65(3), pp.674–685.

18. Wesolowska-Andersen, A., Bahl, M.I., Carvalho, V., Kristiansen, K., Sicheritz-Pontén, T., Gupta, R. and Licht, T.R., 2014. Choice of bacterial DNA extraction method from fecal material influences community structure as evaluated by metagenomic analysis. Microbiome, 2(1), pp.1–11.

19. Bjerre, R.D., Hugerth, L.W., Boulund, F., Seifert, M., Johansen, J.D. and Engstrand, L., 2019. Effects of sampling strategy and DNA extraction on human skin microbiome investigations. Scientific reports, 9(1), pp.1–11.

20. Chen, J., Yue, Y., Wang, L., Deng, Z., Yuan, Y., Zhao, M., Yuan, Z., Tan, C. and Cao, Y., 2020. Altered gut microbiota correlated with systemic inflammation in children with Kawasaki disease. Scientific Reports, 10(1), pp.1–12.

21. Zhang, F., Yue, L., Fang, X., Wang, G., Li, C., Sun, X., Jia, X., Yang, J., Song, J., Zhang, Y. and Guo, C., 2020. Altered gut microbiota in Parkinson’s disease patients/healthy spouses and its association with clinical features. Parkinsonism & Related Disorders, 81, pp.84–88.

22. Amara, A. A., and A. Shibl. 2015. Role of Probiotics in health improvement, infection control and disease treatment and management.” Saudi pharmaceutical journal, 23.2, pp.107–114.

23. Sinha, R., Abu-Ali, G., Vogtmann, E., Fodor, A.A., Ren, B., Amir, A., Schwager, E., Crabtree, J., Ma, S., Abnet, C.C. and Knight, R., 2017. Assessment of variation in microbial community amplicon sequencing by the Microbiome Quality Control (MBQC) project consortium. Nature biotechnology, 35(11), pp.1077–1086.

24. Costea, P.I., Zeller, G., Sunagawa, S., Pelletier, E., Alberti, A., Levenez, F., Tramontano, M., Driessen, M., Hercog, R., Jung, F.E. and Kultima, J.R., 2017. Towards standards for human fecal sample processing in metagenomic studies. Nature biotechnology, 35(11), pp.1069–1076.

25. Heikema, A.P., Horst-Kreft, D., Boers, S.A., Jansen, R., Hiltemann, S.D., de Koning, W., Kraaij, R., de Ridder, M.A., van Houten, C.B., Bont, L.J. and Stubbs, A.P., 2020. Comparison of Illumina versus nanopore 16S rRNA gene sequencing of the human nasal microbiota. Genes, 11(9), p.1105.

26. Bag, S., Saha, B., Mehta, O., Anbumani, D., Kumar, N., Dayal, M., Pant, A., Kumar, P., Saxena, S., Allin, K.H. and Hansen, T., 2016. An improved method for high quality metagenomics DNA extraction from human and environmental samples. Scientific reports, 6(1), pp.1–9.

27. Luo, M., Liu, Y., Wu, P., Luo, D.X., Sun, Q., Zheng, H., Hu, R., Pandol, S.J., Li, Q.F., Han, Y.P. and Zeng, Y., 2017. Alternation of gut microbiota in patients with pulmonary tuberculosis. Frontiers in physiology, 8, pp.822.

28. Hu, Y., Feng, Y., Wu, J., Liu, F., Zhang, Z., Hao, Y., Liang, S., Li, B., Li, J., Lv, N. and Xu, Y., 2019. The gut microbiome signatures discriminate healthy from pulmonary tuberculosis patients. Frontiers in cellular and infection microbiology, 9, pp.90.

29. Wang, S., Yang, L., Hu, H., Lv, L., Ji, Z., Zhao, Y., Zhang, H., Xu, M., Fang, R., Zheng, L. and Ding, C., 2021. Characteristic gut microbiota and metabolic changes in patients with pulmonary tuberculosis. Microbial biotechnology. 0(0), pp.1–14.

30. Shi, W., Hu, Y., Ning, Z., Xia, F., Wu, M., Hu, Y.O., Chen, C., Prast-Nielsen, S. and Xu, B., 2021. Alterations of gut microbiota in patients with active pulmonary tuberculosis in China: a pilot study. International Journal of Infectious Diseases, 111, pp.313–321.

31. Li, W., Zhu, Y., Liao, Q., Wang, Z. and Wan, C., 2019. Characterization of gut microbiota in children with pulmonary tuberculosis. BMC pediatrics, 19(1), pp.1–10.

32. Khaliq, A., Ravindran, R., Afzal, S., Jena, P.K., Akhtar, M.W., Ambreen, A., Wan, Y.J.Y., Malik, K.A., Irfan, M. and Khan, I.H., 2021. Gut microbiome dysbiosis and correlation with blood biomarkers in active-tuberculosis in endemic setting. PloS one, 16(1), p.e0245534.

33. Hart, M.L., Meyer, A., Johnson, P.J. and Ericsson, A.C., 2015. Comparative evaluation of DNA extraction methods from feces of multiple host species for downstream next-generation sequencing. PloS one, 10(11), p.e0143334.

34. Kumar, J., Kumar, M., Gupta, S., Ahmed, V., Bhambi, M., Pandey, R. and Chauhan, N.S., 2016. An improved methodology to overcome key issues in human fecal metagenomic DNA extraction. Genomics, proteomics & bioinformatics, 14(6), pp.371–378.

35. Yuan, S., Cohen, D.B., Ravel, J., Abdo, Z. and Forney, L.J., 2012. Evaluation of methods for the extraction and purification of DNA from the human microbiome. PloS one, 7(3), p.e33865.

36. Phillips, D.C., 1967. The hen egg-white lysozyme molecule. Proceedings of the National Academy of Sciences of the United States of America, pp.483–495.

37. Browder, H.P., Zygmunt, W.A., Young, J.R. and Tavormina, P.A., 1965. Lysostaphin: enzymatic mode of action. Biochemical and biophysical research communications, 19(3), pp.383–389.

38. Schindler, C.A. and Schuhardt, V., 1964. Lysostaphin: a new bacteriolytic agent for the Staphylococcus. Proceedings of the National Academy of Sciences of the United States of America, 51(3), pp.414.

39. Clarke, A.J. and Dupont, C., 1992. O-acetylated peptidoglycan: its occurrence, pathobiological significance, and biosynthesis. Canadian journal of microbiology, 38(2), pp.85–91.

40. Zipperle Jr, G.F., Ezzell Jr, J.W. and Doyle, R.J., 1984. Glucosamine substitution and muramidase susceptibility in Bacillus anthracis. Canadian journal of microbiology, 30(5), pp.553–559.

41. Ariefdjohan, M.W., Savaiano, D.A. and Nakatsu, C.H., 2010. Comparison of DNA extraction kits for PCR-DGGE analysis of human intestinal microbial communities from fecal specimens. Nutrition journal, 9(1), pp.1–8.

42. Li, F., Hullar, M.A. and Lampe, J.W., 2007. Optimization of terminal restriction fragment polymorphism (TRFLP) analysis of human gut microbiota. Journal of microbiological methods, 68(2), pp.303–311.

43. Nylund, L., Heilig, H.G., Salminen, S., de Vos, W.M. and Satokari, R., 2010. Semi-automated extraction of microbial DNA from feces for qPCR and phylogenetic microarray analysis. Journal of microbiological methods, 83(2), pp.231–235.

44. Lim, M.Y., Song, E.J., Kim, S.H., Lee, J. and Nam, Y.D., 2018. Comparison of DNA extraction methods for human gut microbial community profiling. Systematic and applied microbiology, 41(2), pp.151–157.

45. Wagner Mackenzie, B., Waite, D.W. and Taylor, M.W., 2015. Evaluating variation in human gut microbiota profiles due to DNA extraction method and inter-subject differences. Frontiers in microbiology, 6, pp.130.

46. Panek, M., Paljetak, H.Č., Barešić, A., Perić, M., Matijašić, M., Lojkić, I., Bender, D.V., Krznarić, Ž. and Verbanac, D., 2018. Methodology challenges in studying human gut microbiota–effects of collection, storage, DNA extraction and next generation sequencing technologies. Scientific reports, 8(1), pp.1–13.

47. Hu, Y., Yang, Q., Liu, B., Dong, J., Sun, L., Zhu, Y., Su, H., Yang, J., Yang, F., Chen, X. and Jin, Q., 2019. Gut microbiota associated with pulmonary tuberculosis and dysbiosis caused by anti-tuberculosis drugs. Journal of Infection, 78(4), pp.317–322.

48. Maji, A., Misra, R., Dhakan, D.B., Gupta, V., Mahato, N.K., Saxena, R., Mittal, P., Thukral, N., Sharma, E., Singh, A. and Virmani, R., 2018. Gut microbiome contributes to impairment of immunity in pulmonary tuberculosis patients by alteration of butyrate and propionate producers. Environmental microbiology, 20(1), pp.402–419.

